## Supplemental Material for "Concurrent gliomas in patients with multiple sclerosis"

### Supplementary Figure 1

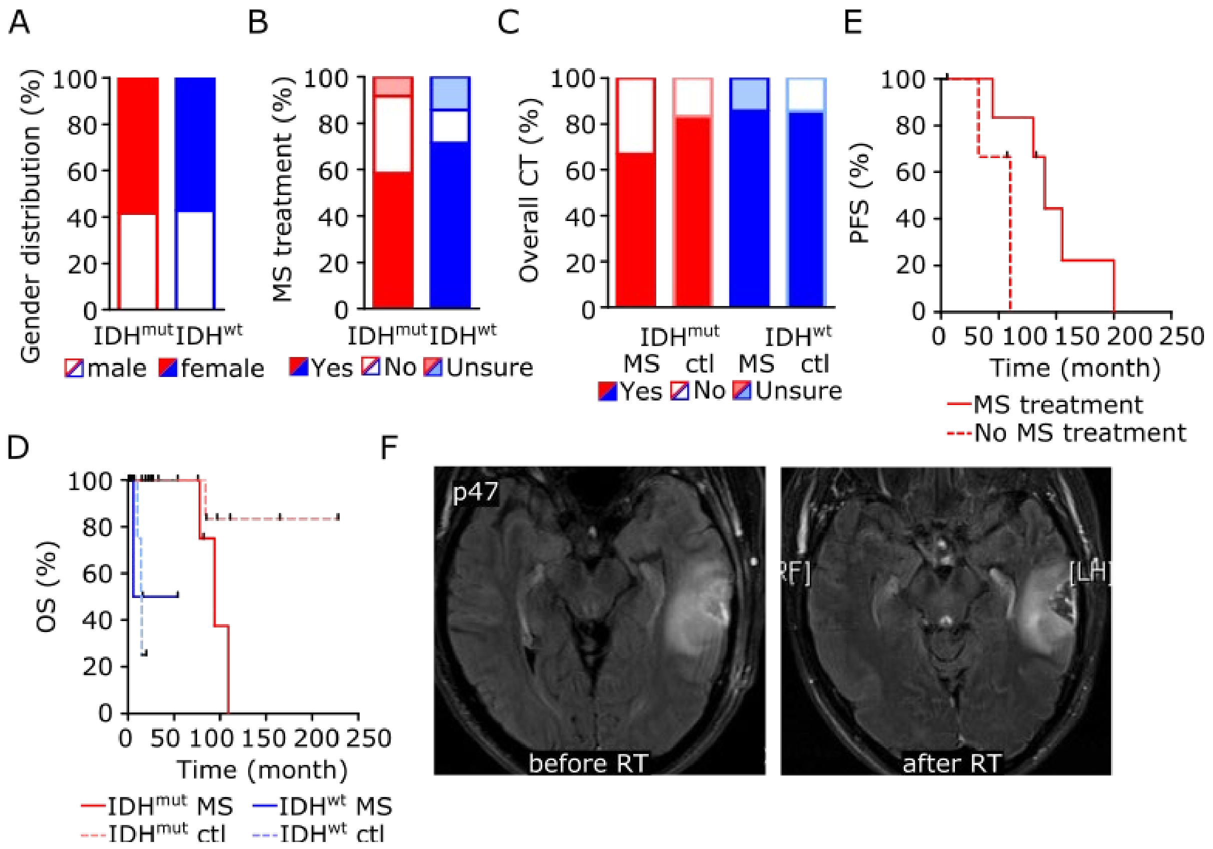

**Clinical and radiological characteristics of 19 patients with multiple sclerosis and concurrent glioma.**

**A** Gender distribution of patient and control cohorts.

**B** Percentage of patients with and without disease-modifying treatment during the course of MS depending on tumoral IDH mutation status.

**C** Percentage of patients with and without chemotherapy (CT) treatment during the course of brain tumor disease depending on concurrent MS diagnosis and tumoral IDH mutation status.

**D** Kaplan–Meier analysis of progression-free survival (PFS) of patients with IDH mutant astrocytoma and concurrent multiple sclerosis after primary tumor treatment depending on immunomodulatory therapy.

**E** Kaplan–Meier analysis of overall survival (OS) after primary tumor treatment depending on IDH status and concurrent multiple sclerosis diagnosis.

**F** MRI with axial FLAIR images of patient p47 showing the tumoral lesion before and after RT with stable disease according to RANO criteria.

**Supplementary Figure 2**

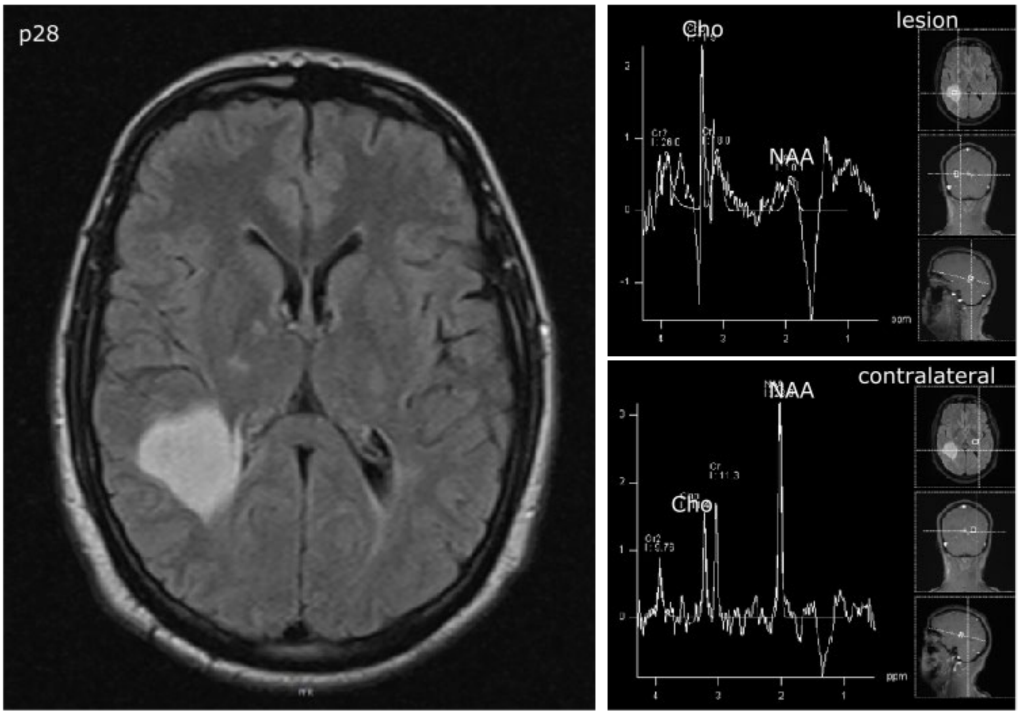

**MRI spectroscopy**

MRI scan of multiple sclerosis patient p28 with axial FLAIR images and spectroscopy demonstrating choline-increase and N-acetyl-aspartate decrease in an IDH-mutant astrocytoma WHO grade II.

**Supplementary Figure 3**

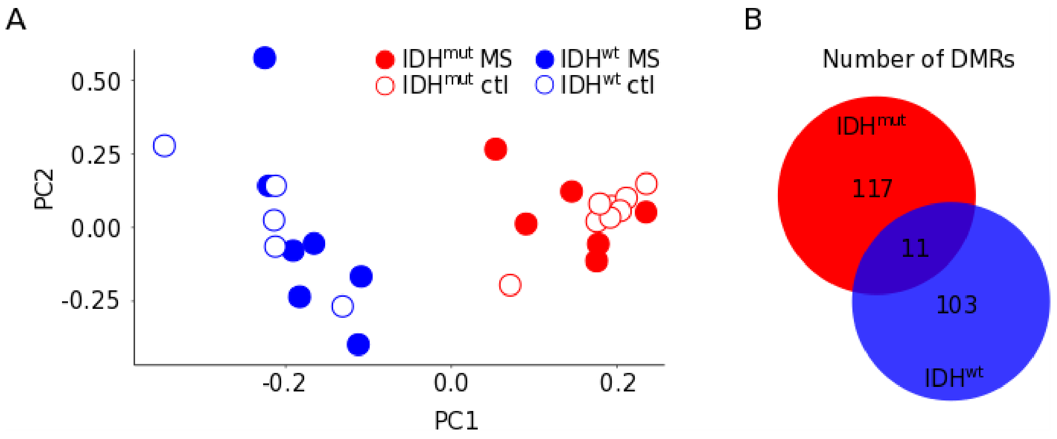

**Genome-wide DNA methylation changes of gliomas in patients with concurrent multiple sclerosis**

**A** Principal component analysis.

**B** Schematic representation of numbers of differentially methylated regions (DMRs) in gliomas compared between patients with concurrent multiple sclerosis and control patients without multiple sclerosis depending on tumoral IDH mutation status.

**Supplementary Table 1 Patient characteristics (n=26)**

| Characteristic | Number of patients (%) |
|---|---|
| Male | 10 (38%) |
| Median age at tumor diagnosis (y) | 42 (15–67) |
| Tumor histology | |
| Astrocytoma | 26 (100%) |
| Oligodendroglioma | 0 (0%) |
| Tumor WHO grade | |
| I | 3 (12%) |
| II | 14 (54%) |
| III | 2 (8%) |
| IV | 7 (27%) |
| IDH1 mutation status | |
| Wildtype | 8 (31%) |
| Mutant | 12 (46%) |
| Unsure | 6 (23%) |
| MGMT promotor status | |
| Hypermethylated | 13 (50%) |
| Hypomethylated | 4 (15%) |
| Unsure | 9 (35%) |
| Tumor location | |
| Frontal | 7 (27%) |
| Temporal | 12 (44%) |
| Parietal | 1 (4%) |
| Occipital | 4 (15%) |
| Brainstem | 1 (4%) |
| Cerebellar | 1 (4%) |
| Surgery | |
| Biopsy | 14 (54%) |
| Partial resection | 2 (8%) |
| Complete resection | 10 (38%) |
| Overall tumor-specific therapy | |
| Radiotherapy | 19 (73%) |
| Chemotherapy | 18 (69%) |
| Median age at MS diagnosis (y) | 33 (16–61) |
| MS symptom course | |
| Relapsing remitting MS | 19 (73%) |
| Primary progressive MS | 0 (0%) |
| Secondary progressive MS | 4 (15%) |
| Inflammatory lesion location | |
| Optic neuritis | 7 (27%) |
| Cerebral | 26 (100%) |
| Spinal | 6 (23%) |
| Overall immunomodulatory therapy | |
| Yes | 15 (58%) |

| | |
|---|---|
| No | 5 (19%) |
| Unsure | 6 (23%) |

**Supplementary Table 2 Glioma risk in patients with multiple sclerosis**

| Study cohort | Total number of patients | Number of patients with MS (%) | |
|---|---|---|---|
| Glioma Heidelberg | 2809 | 12 (0.4) | **Odds ratio (95% CI)** |
| Glioma CPRD* | 3112 | 9 (0.3) | 1.48 (0.62 – 3.52) |
| Control CPRD* | 31120 | 85 (0.3) | 1.57 (0.86 – 2.87) |

Glioma risk among MS patients of the Glioma Heidelberg cohort was compared to risks of Glioma CPRD and Control CPRD cohorts previously published (*(Anssar *et al.*, 2020); 1:10 matched case-control study within the CPRD (Clinical Practice Research Datalink)).

**Supplementary Table 3 Characteristics of patients with IDH1-wildtype glioblastoma WHO grade IV**

| Characteristic | Number of MS patients (%) n=7 (100%) | Number of ctl patients (%) n=7 (100%) |
|---|---|---|
| Male | 3 (43%) | 4 (57%) |
| Median age at tumor diagnosis (y) | 59 (42–67) | 59 (41–71) |
| Glioblastoma manifestation | | |
| New symptoms | 6 (86%) | 6 (86%) |
| MRI incidental finding | 0 (0%) | 1 (14%) |
| Unsure | 1 (14%) | 0 (0%) |
| Tumor location | | |
| Frontal | 2 (29%) | 2 (29%) |
| Temporal | 2 (29%) | 1 (14%) |
| Parietal | 0 (0%) | 2 (29%) |
| Occipital | 3 (43%) | 2 (29%) |
| Brainstem | 0 (0%) | 0 (0%) |
| Cerebellar | 0 (0%) | 0 (0%) |
| Surgery | | |
| Biopsy | 2 (29%) | 1 (14%) |
| Partial resection | 1 (14%) | 4 (57%) |
| Complete resection | 4 (57%) | 2 (29%) |
| MGMT promotor status | | |
| Hypermethylated | 1 (14%) | 2 (29%) |
| Hypomethylated | 4 (57%) | 4 (57%) |
| Unsure | 2 (29%) | 1 (14%) |
| 1st treatment | | |
| Best supportive care | 0 (0%) | 1 (14%) |
| Radiotherapy only | 1 (14%) | 0 (0%) |
| Chemotherapy only | 1 (14%) | 0 (0%) |
| Radiochemotherapy | 5 (71%) | 6 (86%) |
| Median PFS (month) | 2.5 | 8 |

**Supplementary Table 4 Characteristics of patients with IDH1-mutant astrocytoma WHO grade II**

| Characteristic | Number of MS patients (%) n=12 (100%) | Number of ctl patients (%) n=12 (100%) |
|---|---|---|
| Male | 5 (42%) | 8 (67%) |
| Median age at tumor diagnosis (y) | 32.5 (21–48) | 31.5 (22–49) |
| Tumor manifestation | | |
| Symptomatic | 4 (33%) | 12 (100%) |
| MRI incidental finding | 7 (58%) | 0 (0%) |
| Unsure | 1 (8%) | 0 (0%) |
| Tumor location | | |
| Frontal | 4 (33%) | 9 (75%) |
| Temporal | 7 (58%) | 3 (25%) |
| Parietal | 0 (0%) | 0 (0%) |
| Occipital | 1 (8%) | 0 (0%) |
| Brainstem | 0 (0%) | 0 (0%) |
| Cerebellar | 0 (0%) | 0 (0%) |
| Surgery | | |
| Biopsy | 6 (50%) | 3 (25%) |
| Partial resection | 1 (8%) | 3 (25%) |
| Complete resection | 5 (42%) | 6 (50%) |
| 1st treatment | | |
| Watch + wait | 7 (58%) | 6 (50%) |
| Radiotherapy | 3 (25%) | 3 (25%) |
| Chemotherapy | 2 (17%) | 5 (42%) |
| Unsure | 1 (8%) | 0 (0%) |
| Median PFS (month) | 32 | 64 |

**Supplementary Table 5: Multiple sclerosis course**

| | IDH-mutant, WHO grade II | IDH-wildtype, WHO grade IV |
|---|---|---|
| Mean age at MS manifestation (y) | 31 (16–47) | unsure |
| Mean age at MS diagnosis (y) | 31 (16–48) | 40 (30–61) |
| Disease sequence | | |
| Glioma first | 4 (33%) | 0 (0%) |
| Simultaneous | 6 (50%) | 0 (0%) |
| MS first | 1 (8%) | 7 (100%) |
| median time between MS and tumor diagnosis (y) | 0 (-7–13) | 13 (1–34) |
| Cerebral inflammatory lesions | 12 (100%) | 7 (100%) |
| Overall immunomodulatory therapy | | |
| Yes | 7 (58%) | 3 (43%) |
| No | 4 (33%) | 1 (14%) |
| Unsure | 1 (8%) | 3 (43%) |

| | | |
|---|---|---|
| Overall RT | 10 (83%) | 5 (71%) |
| MS progression in the 12 months preceding RT | 0 (0%) | 0 (0%) |
| MS progression in the 12 months after RT | 5 (42%)*, ** | 1 (20%)* |
| Overall CT | 8 (67%) | 6 (86%) |
| MS progression in the 12 months preceding CT | 0 (0%) | 0 (0%) |
| MS progression in the 12 months after CT | 1 (12%)* | 1 (17%)* |

* combined RCT, ** three (60%) with immunomodulatory treatment

**Supplementary Table 6 Common differentially methylated regions (DMR)**

| Genetic region | Annotated gene | Immune-related function |
|---|---|---|
| 3q24 | ZIC4 | |
| 4q23 | EIF4E | Regulation of immune function |
| 5q31.3 | TMCO6 | Interleukin region |
| 5q31.3 | PCDHB3 | Interleukin region |
| 5q35.1 | DUSP1 | Autoimmune disease risk |
| 5p15.33 | C5orf38 | |
| 6p22.1 | RNF39 | HLA region |
| 6p21.32 | PRRT1 | HLA region |
| 10p12.1 | MKX | |
| 12p13.1 | GPR19 | |
| 18p11.21 | CIDEA | |
